# Genetic, Hormonal, and Psychosocial Factors Underly Sex Differences in Psychotic-Like Experience Trajectories in Adolescence

**DOI:** 10.64898/2026.11.16.26368188

**Authors:** Cara Ly, Chaoyu Liu, Julia Ciarcia, Seoyeon Lee, Keira J. A. Johnston, Adrianna P. Kępińska, Laura M. Huckins

## Abstract

Psychotic-like experiences (PLEs) are common during adolescence (prevalence: 17-60%), however, sex differences in the biopsychosocial factors associated with PLEs, particularly pubertal development and sex hormones, and PLE trajectories remains less understood. Here, we leverage data from the Adolescent Brain Cognitive Development (ABCD) Study to examine genetic, psychosocial, and hormonal correlates of PLE risk and persistence in 7,870 adolescents reporting at least one PLE across baseline to year 3 follow-up. Among individuals of European ancestry, higher genetic risk for schizophrenia was positively associated with PLEs. Sex stratified multivariate linear regression analyses revealed sex-specific differences in associations between social factors and the outcome, with parent acceptance (β = -0.09, p = 2.47×10^−02^ in males; β = -0.44, p = 1.5×10^−20^ in females) and overt aggression (β = 0.038, p = 1.67×10^−02^ in males; β = -0.081, p = 1.53×10^−05^ in females) showing differing directions and magnitudes of effect in males and females. Pubertal development in males and females and age of menarche are associated with PLE risk, while mediation analyses identified estradiol in females as a mediator of the association between puberty and PLEs in premenarchal females. Lastly, multivariate logistic regression analyses and Analysis of Variance showed that higher exposure to psychosocial risk factors, such as discrimination (OR = 1.47, p = 1.11×10^−04^) and social victimization (OR = 1.05, p = 2.02×10^−02^), earlier pubertal development, and earlier age of menarche predict membership to the persistent PLE trajectory. These findings highlight sex- specific social correlates of adolescent PLEs and implicate pubertal timing and hormonal pathways in PLE risk and persistence. Understanding how genetic, social, and biological factors jointly shape PLE trajectories may help inform early identification of psychosis risk.

## INTRODUCTION

Psychotic-like experiences (PLEs) encompass auditory and visual hallucinations, delusions, disorganized thoughts and speech, and paranoia. PLEs can occur in individuals who do not meet diagnostic criteria for a psychotic disorder. While 5-7%^1^ of the general population report having PLEs, they are relatively common during adolescence, with approximately 14-17%^2–4^, and in some cases up to more than 60%^1,5^, of adolescents reporting such experiences. The accumulation of PLEs during adolescence elevates the risk for psychosis^6–8^ and other psychiatric disorders such as schizophrenia, anxiety, and major depressive disorder^3,9,10^, and thus, PLEs represent a potential indicator of psychiatric vulnerability.

Like other psychiatric traits, the onset of PLEs in adolescence is explained both by genetics and environment. Twin studies have established significant heritability (15-59%^11,12^) with varying heritability for specific symptoms, such as hallucination and paranoia. SNP heritability from genome-wide association studies (GWAS) is relatively low (3-9%^8,12^), suggesting an impact of environmental factors on PLEs. Previous studies have identified environmental factors that increase the risk for PLEs in adolescence, including socioeconomic status and urbanization^13,14^ with individuals being ∼80% more likely to develop PLEs living in urban neighborhoods^15^.

Stress, trauma, and social victimization have also been associated with increased PLE risk, with adolescents who report at least one traumatic experience being 2-3 times more likely experience PLEs^16–21^. One study identified adolescents with PLEs being 4-6 times more likely to experience child abuse or sexual abuse. Moreover, increasing the number of traumatic events experiences has been associated with increased number, frequency, and severity of PLEs^18,19^. However, it remains unclear to what extent these biopsychosocial factors jointly contribute to PLE risk, and whether their effects differ between males and females.

Prevalence and symptomatology of PLEs differ by sex, with males more commonly exhibiting negative symptoms, such as social withdrawal and apathy, and females more commonly exhibiting affective symptoms, such as depression and anxiety. These sex differences mirror sex differences observed among adults with psychosis and schizophrenia, with males typically showing an earlier age of onset (19–25 years) than females, who exhibit peaks in onset during the mid-20s and again in midlife (45–55 years)^22,23^. These differences are unlikely to be caused by a single factor and may reflect the interaction between biological, developmental, and environmental influences. From a biological perspective, evidence from epidemiological and functional studies supports a role for gonadal hormones contributing to sex differences in vulnerability to PLEs and schizophrenia. In adults, lower estrogen levels in females^24–27^ and lower testosterone levels in males^28–30^ have been associated with increased psychosis and schizophrenia risk. These hormonal influences may be particularly relevant during adolescence, when substantial changes in gonadal hormone levels accompany pubertal development. Previous studies have shown that earlier age at menarche is associated with an increased risk of PLEs, suggesting that the timing of pubertal development may contribute to vulnerability to PLEs during adolescence^31–33^. However, we do not yet know whether the relationship between pubertal development and PLEs differs by sex and whether changes in sex hormones during puberty contribute to these differences.

Previous longitudinal studies have demonstrated that PLEs are not static during adolescence, but instead follow distinct developmental trajectories which vary in onset and progression over time^34^. In previous studies, 80% of adolescents report having few or decreasing PLEs, however, as puberty progresses, a subset of adolescents experience persisting or increasing PLEs^35^. Studies have identified several distinct PLE trajectories, including low or stable, decreasing, increasing, and persistent trajectories, the latter characterized by fluctuating PLEs that occur with greater frequency over time, highlighting substantial individual differences in the developmental course of these experiences^36^. Importantly, individuals with persistent or increasing PLEs have been shown to experience a higher burden of psychiatric disorders compared with those whose PLEs decrease over time^37,38^. While studies have identified psychosocial factors, such as trauma and socioeconomic adversity, that contribute to persisting PLEs, studies have not yet explored sex differences and a wide variety of psychosocial factors in predicting trajectory membership.

Understanding how biological, developmental, and environmental factors contribute to sex differences in PLEs may provide insight into the mechanisms underlying vulnerability to psychopathology. We investigated the contributions of genetic, psychosocial, and developmental factors to the risk and persistence of PLEs during adolescence using data from the Adolescent Brain Cognitive Development (ABCD) Study, a large, longitudinal cohort of approximately 11,800 adolescents with extensive phenotypic, psychosocial, genetic, and salivary hormone data. First, we examined genetic liability for schizophrenia and identified psychosocial factors associated with increased PLE risk. Second, we investigated whether pubertal development and age at menarche were associated with PLE risk and whether sex hormones mediated the association between pubertal development and PLEs. Finally, we identified longitudinal PLE trajectories and examined the genetic, psychosocial, and developmental factors associated with trajectory membership.

## METHODS

### 1. Participants

The Adolescent Brain Cognitive Development Study (ABCD Study) release 5.1 includes longitudinal data from 11,875 participants who were recruited between ages 9 and 10 years at baseline (BL) across 21 research sites throughout the United States. Details of the ABCD Study recruitment strategy, protocols, and design can be found elsewhere^39,40^. We used participants from BL, and follow-up years 1 (Y1), 2 (Y2), and 3 (Y3). Follow-up year 4 was excluded due to many participants missing data, resulting in less power in analyses. Participants with missing PLE scores, environmental exposure, puberty and hormone data across the four time points were excluded from analyses.

### 2. Assessment Measures Psychotic-Like Experiences

The ABCD study uses the Prodromal Questionnaire-Brief Child Version (PQ-BC) to assess PLEs in adolescents. The PQ-BC is 21-item child-reported questionnaire asking participants whether they have experienced various types of PLEs. Types of PLEs include visual and auditory hallucinations, delusions, disorganized thoughts and speech, and paranoia. The PQ-BC includes two scores; 1) total PLE score: the sum of yes to each question, ranging from 0-21, and 2) the distress score: sum of responses to how distressed participant is for each question, ranging from 0-126^41^. Total PLE scores were used for all analyses, given the absence of significant differences in findings when distressing PLE scores were examined. Participants were included in this study if they had a PLE score of 0-21 across each timepoint. Within the cohort chosen for this study (N = 11,857), 7,273 participants reported having at least one PLE at BL, decreasing to 5,536, 4,761, and 3,994 at the Y1, Y2, and Y3 follow-ups, respectively due to participant drop out.

#### Sex

For the purposes of this study, sex was defined using parent responses in the demographics questionnaire.

#### Psychosocial Factors

The ABCD Study contains a comprehensive set of environmental questionnaires capturing diverse domains, including sociodemographic characteristics, social stressors, and other psychosocial factors **(Table S1)**. Sociodemographic factors used in this study were assessed in the Demographics questionnaire where parents/caregivers were asked about the age and sex of the child, household income, and experiences due to low-income. The low-income experiences score was derived from a sum of seven low-income experience questions from the demographic questionnaire, ranging from 0-7. Parent acceptance was calculated from summary scores in the youth-reported Children’s Report of Parental Behavioral Inventory which asks children about their perception of their parent/caregiver’s love and acceptance towards them. The parent score and caregiver scores are the means of the answer to five questions, both with a score ranging from 1 (“Not like him/her) to 3 (“A lot like him/her”).

Adversity and social stressors were assessed through various questionnaires. The Family Conflict questionnaire evaluates aspects of the home environment (e.g., yelling, fighting, criticism). A total family conflict score, ranging from 0–9, was calculated as the sum of “True” responses to the nine questionnaire items. Parent/caregiver neglect was assessed using the Multidimensional Neglectful Behavior Scale, which captures youth-reported experiences of neglectful parental or caregiver behaviors. A parent neglect score was derived by averaging responses to 10 questionnaire items evaluating parent/caregiver attentiveness to the youth’s school, safety, and behavioral concerns. Responses were rated on a 4-point scale ranging from 0 (“Never”) to 3 (“A lot”). Bullying, including both victimization and perpetration, we measured using the Peer Experiences questionnaire which assess youth-reported experiences with bullying as a victim and/or aggressor. The Discrimination Measure is a 7-item questionnaire that asks youth how often they experience discrimination and feelings about Americans. The discrimination measure score is the mean of responses to each question, ranging from 1 (“Almost never”) to 5 (“Very often”).

The youth-reported Life Events questionnaire includes 25 items asking whether the participant has experienced a specific event at any time or in the past year, whether it was a good or bad event, and how much they were affected by the event. Five scores were used from the questionnaire; 1) The life events score – sum of “yes” answers to each question, 2) the bad life events score – sum of life events perceived as bad, 3) the good life events score – sum of life events perceived as good, 4) effected by bad life events score – mean of how much participant was affected by bad life events ranging from 0 (“Not at all”) to 3 (“A lot”). Four scores were derived from the Life Events questionnaire. The parent-related life events score was calculated as the sum of “yes” responses to items concerning parents or caregivers (e.g., “One of the parents/caregivers went to jail?”). The family and friend-related life events score reflected the sum of affirmative responses to events involving family members or friends (e.g., “Family member had a drug and/or alcohol problem?”). The new environment score represented the sum of “yes” responses to experiences involving environmental changes (e.g., “Attended a new school?”). Finally, the personal life events score was calculated as the sum of affirmative responses to events directly experienced by the participant (e.g., “Was a victim of crime, violence, or assault?”).

School environment was assessed with the youth-reported School Risk & Protective Factors questionnaire. The school environment score is the sum of items asking how supported, engaged, and safe the participant feels in their school environment, ranging from 0-6. The school involvement score, ranging from 0-4, is the sum of four items asking participants academic self- confidence, classroom participation, and positive attitude towards school. The school disengaging score, ranging from 0-2, is the sum of two items gauging how important school and grades are to the participant. Participants’ responses to stress were assessed using Will’s Problem-Solving Scale (WPS) and the Barkley Deficits in Executive Functioning Scale (BDEFS). The WPS score was calculated as the sum of six items assessing participants’ problem-solving approaches. The BDEFS assesses executive functioning difficulties, including emotional regulation, impulse control, and self-management. Both the total BDEFS score (sum of all items; range = 14–56) and the symptom summary score (sum of items scored > 3) were used in analyses.

#### Pubertal Development

The pubertal development scale (PDS) ^31,42^ provides a measure of the participant’s perceived physical maturation with questions asking about pubertal characteristics, such as growth in height, growth in body hair, skin changes (pimples), and sex-specific factors including breast growth, menstruation, and voice change. Pubertal development was assessed separately for males and females using sex-specific Pubertal Development Scale (PDS) scores, which range from 1 (prepubertal) to 5 (post pubertal). For males, PDS scores were derived from items assessing body hair growth, voice deepening, and facial hair growth, with summed scores categorized into pre-, early, mid-, late-, and post-pubertal stages based on established thresholds. For females, PDS scores were calculated using items assessing body hair growth and breast development, in conjunction with menarche status, with summed scores similarly categorized into pre-, early, mid-, late-, and post-pubertal stages according to established thresholds.

#### Hormones

The ABCD Study used salivary sampling as a noninvasive method for assessing concentrations of three gonadal hormones - Testosterone, estradiol, and Dehydroepiandrosterone (DHEA) - associated with pubertal development. Details of the saliva collection can be found elsewhere^31^. Testosterone, estradiol, and DHEA were collected from females and only testosterone and DHEA in males. To obtain hormone measures for each participant across all time points (BY through Y3 follow-up), quality control procedures were implemented. Participants were retained if the sex reported at collection matched the sample tube cap color and if no concerns regarding sample quality were noted. Participants with hormone concentrations below assay sensitivity thresholds or with one or more sample quality concerns were excluded. Each sex hormone was measured twice for each participant. When the two values were available, the mean value was used; if only one valid measurement was available, that value was retained.

### 3. Data Analysis

#### Schizophrenia genetic risk among the ABCD cohort

All statistical analyses were conducted using R version 4.4.1. Polygenic risk scores (PRS) were calculated using PRS-CS, a Bayesian polygenic prediction method that incorporates genome- wide association study (GWAS) summary statistics and linkage disequilibrium reference panels. Ancestry-specific summary statistics from the Psychiatric Genomics Consortium Schizophrenia GWAS were used for European, African, American, and East Asian ancestry groups to calculate PRS values.

#### Psychosocial factors associated with PLEs in adolescents

Multivariate linear regressions were performed across four time points, BL – Y3 follow-up, to identify environmental factors that increase or decrease PLE risk. With PLE scores as the outcome, predictors included all environmental factors (see *Psychosocial Factors*) with sex as a covariate. A Bonferroni correction was applied to account for multiple comparisons, resulting in a corrected significance threshold of *p* < 0.0125 (α = 0.05/4). Sex stratified multivariate linear regressions were performed to identify sex differences in associations between PLEs and psychosocial factors in males and females, followed by permutation tests to assess sex differences in effect. Permutation tests were conducted by repeating the linear regression analysis 10,000 times with randomly permuted sex assignments, with p-values calculated as the proportion of permutations yielding effect sizes equal to or more extreme than the observed effect size.

#### Pubertal development association with PLEs and sex hormone mediation

Univariate linear regression analyses were used to examine the association between pubertal development stage, as measured by the PDS score, and PLE risk. Correlation tests were performed to identify the association between age of menarche and age of PLE onset. Mediation analyses were performed using the *Mediation* package (Version 4.5.0) in R. Male and females were separated, and females were further separated into pre-menarche and menstruating due to differences in hormone level differences during the menstrual cycle and excluded females with hormonal contraceptives. Average causal mediation effects (ACMEs) were estimated to quantify the indirect effect of pubertal development on PLEs through hormone levels (testosterone, estradiol, and DHEA), representing the portion of the association between puberty and PLEs that operates through each hormonal mediator (**Figure 3**). Mediation analyses were conducted for BL and Y1 and Y2 follow-up, however, it was not performed in Y3 follow-up due to little variation in pubertal status in that year. A Bonferroni correction was applied to account for multiple comparisons, resulting in a corrected significance threshold of *p* < 0.0056 (α = 0.05/9).

**Figure 1.**
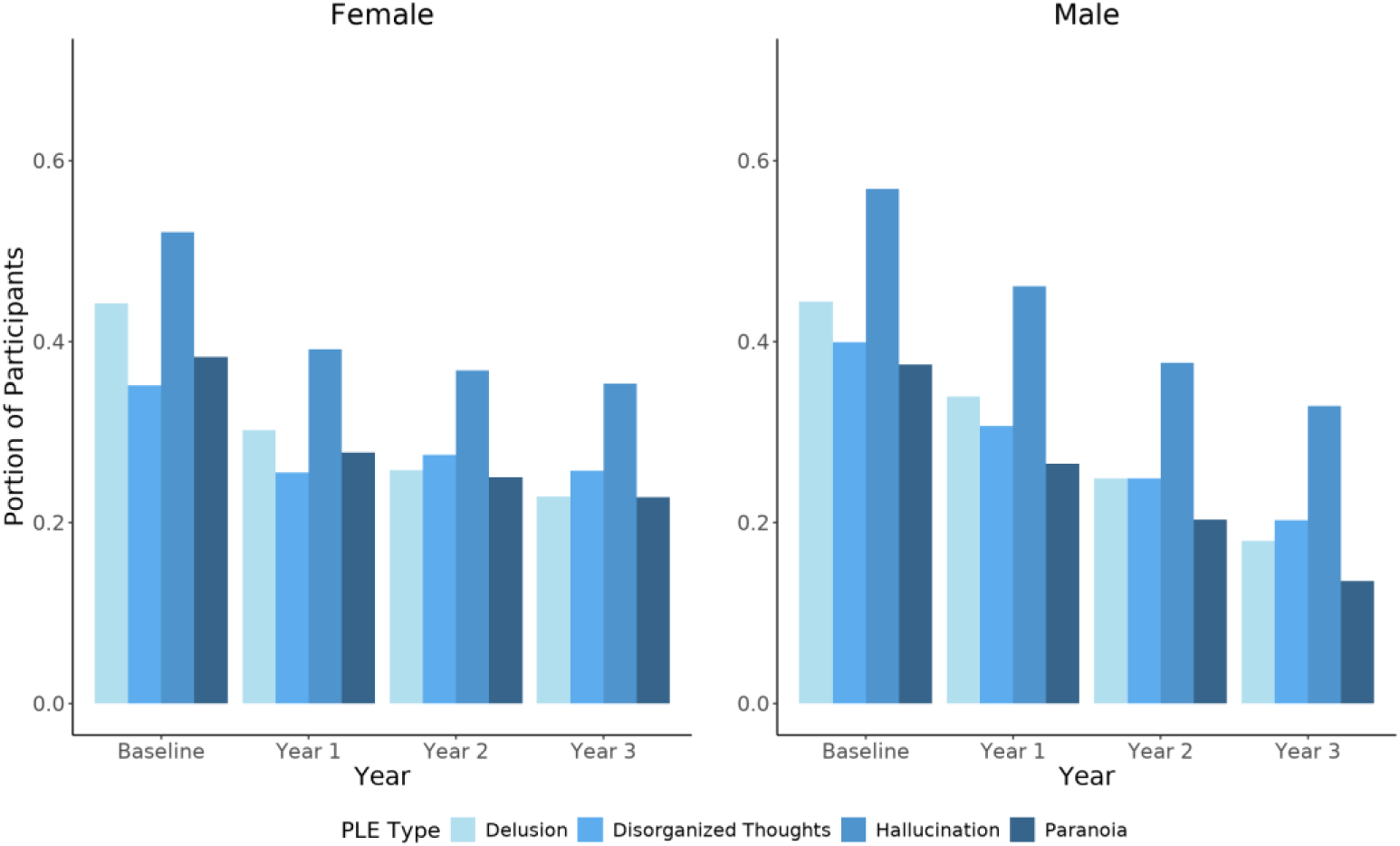
Portion of male and female participants reporting each PLE symptom across study years. Participants reporting at least one symptom were included. Among these participants, there were 6,181 males and 5,673 females at baseline, 5,862 males and 5,348 females at Year 1, 5,735 males and 5,203 females at Year 2, and 5,409 males and 4,896 females at Year 3.

**Figure 2.**
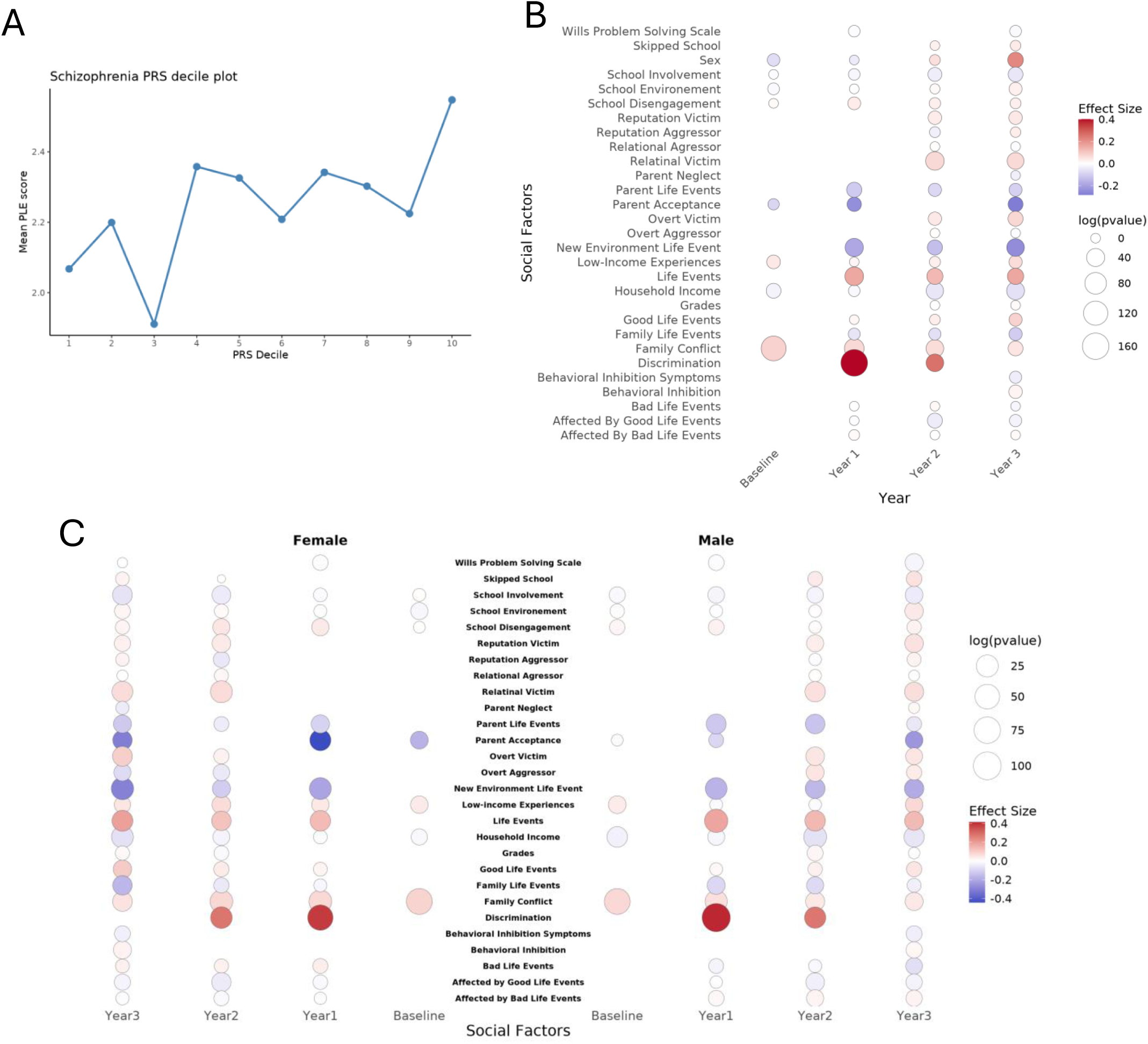
Genetic and environmental contributions to PLE risk in adolescence. Participants reporting at least one PLE were included for every year. (A) European participants within ABCD cohort were separated into PRS deciles and compared to mean PLE scores of participants within that decile. (B) Multivariate linear regression shows psychosocial factors associated with increased PLEs across each year. (C) Sex differences in psychosocial factors that contribute to increased PLEs in adolescents.

**Figure 3.**
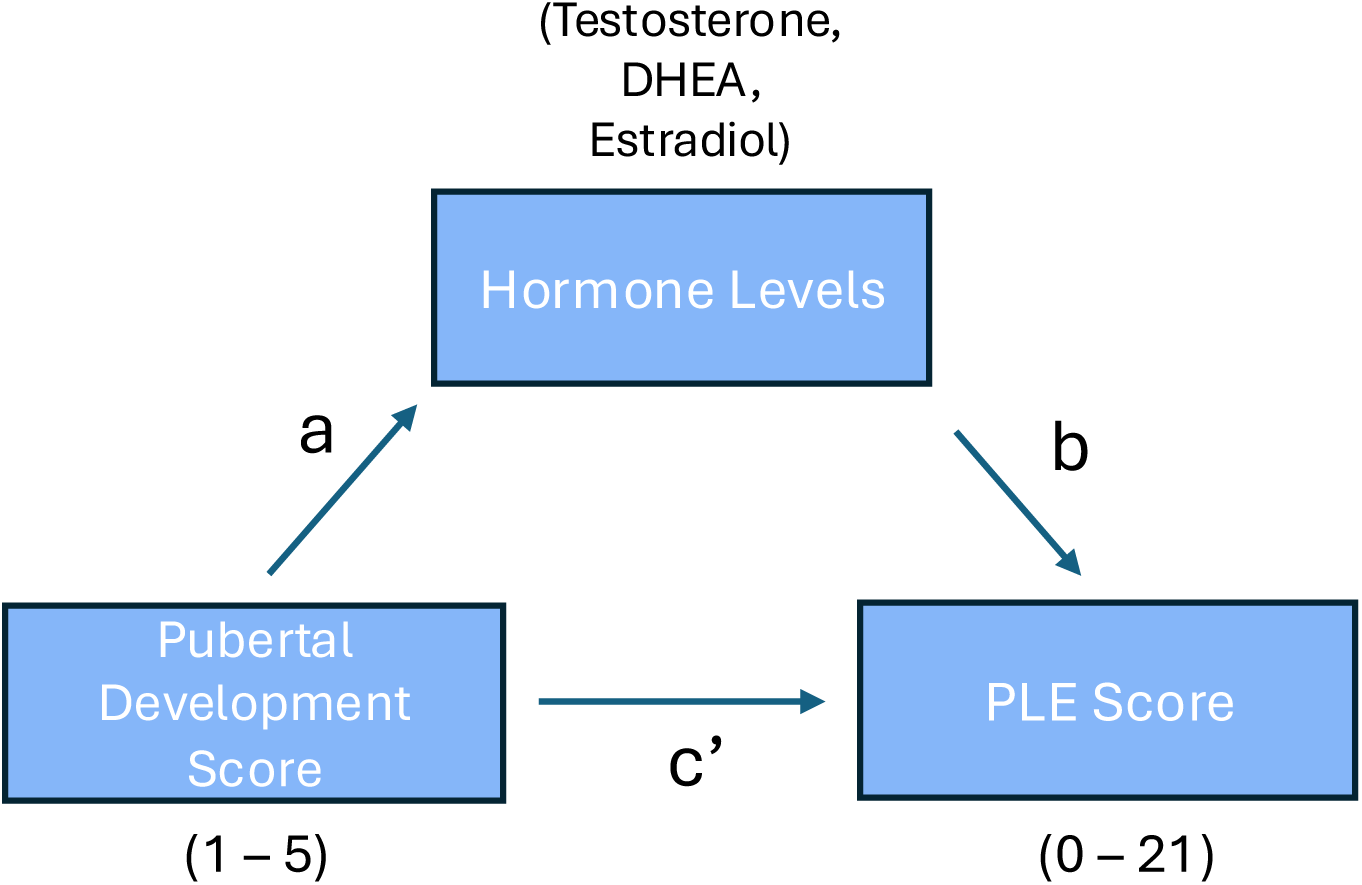
Statistical mediation model illustrating paths a (effect of PDS on hormone levels), b (effect of hormone levels on PLE score controlling for PDS), and c’ (direct effect of PDS on PLE score controlling for hormone levels). Hormones include testosterone, estradiol, and DHEA, pubertal development score (PDS) ranges from 1-5, and PLE score ranges from 0-21.

#### PLE trajectories

PLE longitudinal trajectories were constructed including individuals with available PLE scores across each year (n = 9926). Trajectory groups were selected based on previous trajectory studies where stable, decreasing, increasing, and persistent or fluctuating trajectories were identified^36,37^.

Participants were assigned to five trajectories: no PLEs (n=2047), increase (n=406), decrease (n=1957), stable (n=2809), and fluctuating (n=2698), with definitions as follows:

**No PLEs:** PLE score of 0 across all years
**Stable:** PLE scores remained relatively consistent across follow-up assessments, defined as identical scores or changes of fewer than 3 points between assessments, provided there was no sustained increase or decrease in symptoms.
**Increasing:** PLE scores demonstrated an overall increase across follow-up assessments. Participants with scores of 0 from BL through Y2 who reported a score greater than 0 at Y3 were also classified as having an increasing trajectory, regardless of the magnitude of the increase.
**Decreasing:** PLE scores demonstrated an overall decrease across follow-up assessments. Participants who reported at least one PLE at BL but had scores of 0 at all subsequent follow-up assessments were also classified as having a decreasing trajectory.
**Fluctuating:** PLE scores exhibited substantial bidirectional changes over time, defined as changes of more than 3 points between consecutive assessments in alternating directions (i.e., an increase followed by a decrease, or a decrease followed by an increase).

#### PLE trajectory association with biopsychosocial factors and psychiatric disorders

Using the *nnet* (Version 7.3.19) and *broom* (Version 1.0.6) R packages, multinomial logistic regression was performed to identify social factors associated with trajectory assignment. The stable trajectory was used as the reference group, comparing environmental factors associated with the decrease, increase, and fluctuating trajectories. Analysis of variance (ANOVA) followed by Tukey’s Honestly Significant Difference (HSD) tests was conducted to assess differences in age of pubertal onset and menarche timing among trajectory groups. Using the *PheWAS* package (version 0.99.6.1), phenome-wide association studies (PheWAS) were conducted to identify associations between PLE score and psychiatric disorders assessed using the Kiddie Schedule for Affective Disorders and Schizophrenia (K-SADS) among trajectory groups. K-SADS in the ABCD Study assesses child and adolescent psychiatric symptoms and psychopathology, including symptoms related to mood, anxiety, attention-deficit/hyperactivity disorder (ADHD), and other psychiatric disorders, and includes both parent- and youth-reported responses. For this study, youth-reported K-SADS measures were used as it prioritizes the participants perspective and may capture internalized symptoms that are less observable by parents or caregivers.

Separating the participants, PheWAS were performed for each trajectory to identify which trajectory has higher psychiatric burden. To account for multiple testing across the 127 phenotypes in the PheWAS, we applied a Bonferroni correction, setting the significance threshold at p < [0.05/127].

## RESULTS

### PLEs within the ABCD cohort

We identified 7,273 participants within the ABCD Study who have experienced at least one PLE, with the number of participants decreasing each follow-up year. Sex differences in the proportion of participants reporting at least one PLE shifted across development. Males comprised a significantly greater proportion of participants reporting PLEs at BL (χ²(1) = 9.624, *p* = 0.0019) and Y1 (χ²(1) = 29.903, *p* = 4.54×10^−8^); however, this pattern reversed at Y2 and 3. In Y3, females comprised a significantly greater proportion of participants reporting PLEs (χ²(1) = 18.045, *p* = 2.158×10^−5^). Items assessing hallucinations, paranoia, and disorganized thoughts and speech were the most frequently endorsed. These included hearing unusual sounds, sensing the presence of an unseen person or entity, believing that others were watching or talking about them, and difficulty expressing thoughts clearly **(Table S2)**. To assess sex differences in the distribution of PLE symptom domains, two-sample Kolmogorov–Smirnov tests were conducted at BL and across Years 1–3 follow-up. Significant sex differences were identified between males and females in the BL, and follow-up years 1 and 3. Specifically, boys exhibited higher levels of hallucinations (BL: *D* = 0.0568, *p* = 3.39 × 10⁻⁵; Y1: *D* = 0.0698, *p* = 7.83 × 10⁻⁸; Y3: *D* = 0.0294, *p* = 6.7 × 10⁻⁴) and disorganized symptoms (BL: *D* = 0.0478, *p* = 5.9 × 10⁻⁴; Y1: *D* = 0.0517, *p* = 1.8 × 10⁻⁴; Y3: *D* = 0.0545, *p* = 6.99 × 10⁻⁵). In contrast, girls exhibited higher levels of delusions and paranoia at later follow-ups, with significant sex differences in delusions at Y1 (*D* = 0.0374, *p* = 1.46 × 10⁻²) and Y3 (*D* = 0.0487, *p* = 4.86 × 10⁻⁴) and in paranoia at Y3 (*D* = 0.0922, *p* = 1.02 × 10⁻¹³). (**Table S3**). The portion of participants reporting each symptom domain generally declined over time in both sexes, with hallucinations remaining the most frequently reported symptom domain across all assessments. Females also showed relatively higher portions of delusions and paranoia at later follow-up years, aligning with the emergence of significant sex differences in these symptom domains over time **(Figure 1).**

### PLE longitudinal trajectories reflect persistence, improvement, or worsening over time

PLEs exhibit both variability and persistence over time. Individuals were assigned to four trajectories: increase (n=406), decrease (n=1957), stable (n=2809), and fluctuating (n=2698). A chi-square test of independence revealed that sex distribution differed significantly across PLE trajectory groups, X^2^(3) = 41.35, *p* = 5.51×10^-9^.

### PLE burden is partially explained by schizophrenia genetic risk

To assess whether common genetic liability for schizophrenia contributes to the onset and persistence of PLEs, we tested the association between schizophrenia PRS and PLE trajectories. Higher schizophrenia PRS in European ancestry individuals was significantly associated with increased PLE risk (*p* = 8.65 × 10⁻⁶), with individuals in the highest Schizophrenia PRS deciles exhibiting the highest mean PLE scores **(Figure 2A)**. These findings indicate that greater genetic liability for schizophrenia was associated with a greater burden of PLEs. The addition of schizophrenia PRS explained an additional 0.08% of the variance in PLE scores beyond the covariates (ΔR² = .0008). No statistically significant associations were observed for East Asian, African, or American ancestry groups.

### Psychosocial factors jointly contribute to PLE risk in adolescents

To identify psychosocial risk and protective factors associated with PLE risk in adolescents, we performed a multivariate linear regression analysis, with PLE score as the outcome variable, psychosocial factors as predictors, and sex included as a covariate. Across multiple years, psychosocial factors including life events (Y1: β = 0.159, *p* = 2.17×10^−49^), discrimination (Y1: β = 0.4, *p* = 1.18×10^−167^), and family conflict (Y1: β = 0.07, *p* = 2.53×10^−60^) were associated with increased PLE risk, whereas parent acceptance (Y1: β = -0.24, *p* = 4.97×10^−15^) and higher household income (Y3: β = -0.06, *p* = 3.62×10^−41^) were associated with decreased PLE risk (**Figure 2B, Table S4**). Males exhibited a higher risk for PLEs at BL and Y1 follow-up, whereas females exhibited a higher risk at Y2 and Y3 follow-up.

To assess sex differences in the direction and magnitude of factors associated with PLE risk in adolescents, a sex-stratified multivariate linear regression was performed (**Figure 2C**). We identified sex-specific differences in associations between social factors and PLE risk, with parent acceptance at Y1 follow-up (β = -0.09, p = 2.47×10^−02^ in males; β = -0.44, p = 1.5×10^−20^ in females), and family life events (β =, p = 1.5×10^−20^ in males; β = -0.169, p = 9.59×10^−12^ in females), good life events (β = 0.0524, p = 1.74×10^−02^ in males; β = 0.106, p = 2.54×10^−08^ in females), and overt aggression (β = 0.038, p = 1.67×10^−02^ in males; β = -0.081, p = 1.53×10^−05^ in females) at Y3 follow-up showing differing directions and magnitudes of effect in males and females. Permutation analyses further demonstrated significant sex differences in the magnitude of the observed effects (**Table S5**).

### Later pubertal development and age of menarche are associated with PLE risk

The shift in sex-specific PLE risk observed at Y2 follow-up (ages 11–13 years) prompted an examination of whether pubertal development was associated with PLE risk. Linear regression models, with pubertal stage as the predictor and PLEs as the outcome, indicated that more advanced pubertal development was associated with increased PLE risk in both males (β = 0.939, p = < 2.0×10^−16^) and females (β = 0.523, p =3.26×10^−16^) (**Table S6**). Because menarche represents a key pubertal milestone unique to females and may help clarify sex-specific developmental effects, we further examined its relationship with PLEs. Age at menarche was positively correlated with age of PLE onset (r = 0.23446, p = 2.19 × 10^-10^) and negatively correlated with the total number of PLEs (r = −0.1836, p < 2.2 × 10^-16^), indicating that later menarche was associated with later onset and fewer overall PLEs in females.

### Estradiol mediates the effect of puberty on PLE risk in a sex dependent manner

To assess whether the relationship between puberty and PLEs is mediated by hormone levels, we conducted mediation analyses in BL to Y2 follow-up. The mediation analysis examined puberty as the predictor, PLEs as the outcome, and sex hormones (testosterone, estradiol, or DHEA) as mediators **(Figure 3)**. Analyses were stratified by sex, with females further separated into premenarchal girls. In premenarchal girls at BL, with DHEA as the mediator, results indicated a significant negative indirect effect (*ACME* = −0.023, 95% CI [−0.046, −0.001], *p* = 0.048), suggesting that DHEA partially attenuated the association between pubertal stage and PLEs (**Table S7)**. In contrast, the direct effect remained positive and significant (*ADE* = 0.61, *p* = <0.0001), indicating that later pubertal stage is associated with more PLEs independent of the mediator. The total effect was also positive and significant (*total effect* = 0.588, *p* = <0.0001), suggesting an overall positive association between puberty and PLEs despite the suppressing (negative) indirect pathway through DHEA. In premenarchal girls at Y2 follow-up, we observed a significant positive indirect (*ACME* = 0.016, 95% CI [8.15×10^−05^, 0.04], *p* = 0.05), direct (*ADE* = 0.316, *p* = <0.0001), and total effect (*total effect* = 0.332, *p* = <0.0001) with estradiol as the mediator, suggesting that pubertal development was associated with PLEs through changes in estradiol levels.

#### Psychosocial exposures, puberty, and psychiatric comorbidities predict PLE trajectory membership

We tested for associations between PLE trajectories and biopsychosocial factors to identify factors that may contribute to improved or worsened outcomes (i.e., increase vs. decrease trajectories). To identify differences in genetic risk among individuals that increase, decrease, stay stable, or fluctuate in PLE score over time, the mean PRS was compared between groups. The difference in mean PRS was not significant across each trajectory group (**Figure 4A**). A multinomial logistic regression was conducted to examine differences in psychosocial exposures across trajectory groups, using the stable group as the reference category (**Figure 4B-C**). Results indicated that multiple psychosocial factors significantly differed across the stable, increasing, decreasing, and fluctuating trajectory groups. Higher exposure to psychosocial risk factors, including discrimination (Y2 Follow-up: OR = 1.47, p = 1.11×10^−04^), social victimization (Y2 Follow-up: victim of relational bullying: OR = 1.05, p = 2.02×10^−02^, victim of reputation bullying: OR = 1.07, p = 2.23×10^−03^, victim of overt bullying: OR = 1.07, p = 8.73×10^−03^), and interpersonal life events (Y3 Follow-up: OR = 1.13, p = 2.89×10^−03^), was associated with greater likelihood of membership in the fluctuating trajectory group. In contrast, higher parental acceptance (Y3 Follow-up: OR = 1.3, p = 3.98×10^−03^) and greater exposure to good life events (Y2: OR = 1.1, p = 4.66×10^−02^) were associated with increased likelihood of membership in the decreasing trajectory group. Conversely, higher household income (Y3 Follow-up: OR = 0.89, p = 3.47×10^−14^) and greater parental acceptance (Y3 Follow-up: OR = 0.79, p = 2.77×10^−02^) were associated with reduced likelihood of membership in the fluctuating group.

**Figure 4.**
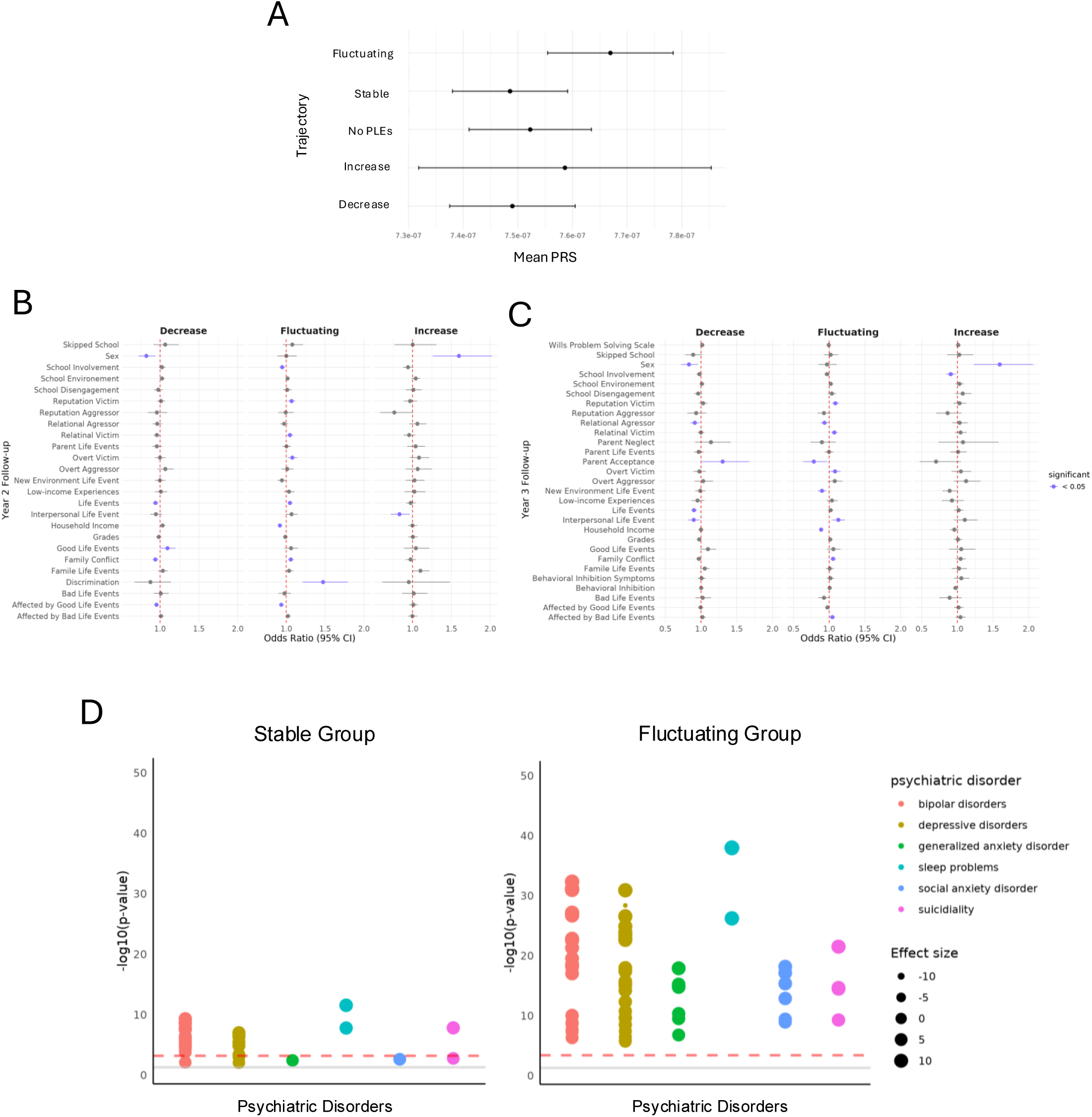
Genetic and environmental contributions to trajectory membership and differences in psychiatric comorbidities among stable and fluctuating trajectories. (A) Mean PRS of all participants in each trajectory. 2047 participants with no PLEs, 1957 in the decrease group, 406 in the increase group, 2809 in the stable group, and 2697 in the fluctuating group. Multinomial logistic regression analysis showing differences in psychosocial factors and trajectory membership with stable as a reference in Year 2 follow-up (B) and Year 3 follow-up (C). (D) Manhattan plot of PheWAS showing the association with PLEs and Psychiatric disorders with the stable and fluctuating group.

A one-way ANOVA was conducted to examine differences in age at pubertal onset across stable, decreasing, increasing, and fluctuating trajectory groups (Table 1). There was a significant effect of trajectory group on pubertal onset, *F*(3, 7701) = 8.266, *p* = 1.73 × 10⁻⁵. Tukey HSD post hoc tests indicated that the fluctuating group had significantly later pubertal onset compared to the decreasing group (mean difference = −0.1644, *p* = 0.0026), as well as compared to both the increasing group (mean difference = 0.2328, *p* = 0.0292) and the stable group (mean difference = 0.1906, *p* = 5×10^-5^). No significant differences were observed between the decreasing, increasing, and stable groups. We further examined age at menarche and observed a significant effect of trajectory group, *F*(3, 2309) = 14.08, *p* = 4.3×10^-9^. Tukey HSD post hoc tests revealed that the fluctuating group experienced significantly earlier menarche compared to the decreasing group (mean difference = −0.2597, *p* < 0.0001), the increasing group (mean difference = −0.2227, *p* = 0.035), and the stable group (mean difference = −0.2762, *p* < 0.0001) **(Table 1)**. In contrast, no significant differences were observed among the decreasing, increasing, and stable groups, suggesting that the effects were primarily driven by earlier menarche in the fluctuating trajectory group.

To assess differences in associations between PLEs and psychiatric disorders across trajectory groups, we conducted a PheWAS within each trajectory using psychiatric symptoms and diagnoses data from the K-SADS questionnaire. PheWAS indicated that the fluctuating group had a higher burden of psychiatric disorders compared to the stable group with there being more symptoms of all six disorders– bipolar, Depressive, generalized anxiety, and social anxiety disorders, sleep problems, and suicidality **(Figure 4D)**. The decrease group also had a higher burden of psychiatric disorders compared to the stable group, similar to the fluctuating group.

The increase group had only two Bonferroni-corrected significant associations with symptoms that were related to major depressive disorder where the participant did not have a continuous period of at least 2 months during which their depressive symptoms were completely absent (β = -0.466, p =1.19×10^−04^).

## DISCUSSION

Our study provides insight into how biopsychosocial factors, such as genetics, hormones, and societal factors, play a role in increasing the risk for PLEs during adolescence using the ABCD Study. We found that liability for schizophrenia is associated with PLEs among European- ancestry adolescents, suggesting that inherited risk contributes to PLE vulnerability. However, genetics represent only one component of the broader biological and environmental factors contributing to PLEs. For example, in line with previous findings, psychosocial factors including discrimination, low socioeconomic status^13^, and traumatic experiences^16,17,19^ all contribute to increased PLE, while parental acceptance decreases likelihood of PLEs.

Within the ABCD cohort, we observed a shift in sex differences in PLE prevalence across development. At BL, males were more likely to report at least one PLE; however, this pattern reversed by follow-up Y3, when females were more likely to report one or more PLEs. We also identified sex differences in PLE symptom profiles, with males reporting more hallucinations and females reporting more delusional and paranoid experiences. The emergence of sex differences in psychopathology is complex and likely reflects the interplay of biological, developmental, social, and environmental factors that change across development. We found that several psychosocial factors were associated with PLE risk differently in male and female adolescents, with differences in both the direction and magnitude of these associations. For example, trauma was more strongly associated with PLE risk among females than males at Follow-up Y3. Interestingly, exhibiting overt aggression was associated with lower PLE risk among females but higher PLE risk among males. Although this finding was unexpected, one possible explanation is that the social dynamics and peer hierarchies of adolescence may differ by sex, such that aggressive behavior may serve different social functions for girls and boys. For example, among girls, aggressive or dominant behaviors may be associated with social status or peer acceptance in certain contexts, potentially reducing exposure to other social stressors that contribute to PLE risk. However, this interpretation should be further investigated to determine mechanisms underlying this sex-specific association.

As previously discussed, sex differences in psychiatric vulnerability are complex and may reflect the influence of multiple biological factors, including pubertal development and sex hormones. Puberty, a crucial developmental stage, is marked by substantial hormonal, physical, and neurodevelopmental changes. Fluctuations in sex hormones during various pubertal stages, with increased testosterone in males and estrogen in females, affect both physical and neurodevelopmental processes. Sex-specific biological changes also occur during puberty, such as menarche. We found that earlier pubertal development and earlier onset of menarche are associated with an increased risk of PLEs. The overall association between pubertal development and PLEs could reflect specific hormones having differing relationships with PLEs.

Our mediation analyses indicate that the association between pubertal development and PLEs differs by sex and hormone levels. Changes in estradiol levels mediated the association between pubertal development and PLEs in premenarchal females. In contrast, DHEA in premenarchal females showed negative indirect effects. Our findings suggest that DHEA in premenarchal females may attenuate the association between pubertal development and PLEs, whereas estradiol may partially account for an increased association. This result was expected as estradiol is known to play a role in the onset of schizophrenia in adult females with lower estradiol levels^23,26,27,43^ and estrogen treatment being used to reduce psychotic symptoms^43^. These findings indicate that hormones represent one pathway through which biological factors contribute to differences in PLE risk across adolescence, along with genetics and psychosocial factors.

We identified four distinct PLE trajectories— increasing, decreasing, stable, and fluctuating— and found that both biological and psychosocial factors were associated with trajectory membership. Multinomial logistic regression analyses indicated that female sex, greater social victimization, lower parental acceptance, and exposure to various forms of trauma were associated with increased odds of membership in the increasing and fluctuating PLE trajectories. These findings were expected as the multivariate regression also showed these psychosocial factors increase the risk of PLEs. Analysis of Variance followed by Tukey’s Honestly Significant Difference post hoc test showed that earlier pubertal development and earlier age at menarche were associated with membership in the fluctuating trajectory compared with the decreasing and stable trajectories. Together, these findings suggest that sex and developmental factors may contribute to differences during PLEs across adolescence, with pubertal timing and age at menarche potentially playing a role in the persistence and fluctuation of PLEs.

There are a few notable limitations of this study. Longitudinal studies tend to experience participant dropout, also leading to missingness within the data. As participants leave the study, the number of individuals available within each trajectory group may decrease, particularly for smaller groups, reducing statistical power and limiting our ability to further define and characterize specific PLE trajectories. Participant attrition may also introduce bias if individuals who discontinue participation differ systematically from those who remain in the study. Second, measures such as the Pubertal Development Questionnaire and KSADS include information reported by both youth and their parents, and responses may differ between informants for the same questions or measures. These discrepancies may reflect differences in how adolescents and parents perceive or report developmental, behavioral, or psychiatric characteristics.

Consequently, reliance on different informants may introduce variability in the measurement of key study variables and potentially influence the results and interpretation of our analyses. Third, the PQ-BC assesses whether a participant has experienced a PLE however, this is based on memory. As a result, a participant may not report a PLE at a given assessment that was also experienced prior to the study period or before the previous assessment. This limitation makes it difficult to determine the precise onset and duration of PLEs and may affect the classification and interpretation of longitudinal PLE trajectories.

In conclusion, we identified sex differences in PLE risk and found that psychosocial adversity, including trauma, low-socioeconomic status, and family conflict, as well as pubertal development were associated with sex differences in PLE burden and longitudinal trajectories. Collectively, these findings suggest that the development and persistence of PLEs may reflect the convergence of genetic susceptibility with social, socioeconomic, and developmental factors.

These findings further highlight adolescence as a critical developmental period for understanding the complex pathways contributing to vulnerability to psychopathology.

## Funding

This work was supported by the Global Talent fund [UKRI2830], the NIMH [MH124839], and NSF [DGE-2139841].

L.M.H. also acknowledges funding from NIMH (R01MH118278, RM1MH132648, R01MH125938) the National Institute of Environmental Health Sciences (R01ES033630), and the Department of Defense.

## Data Availability

All data produced in the present work are contained in the manuscript

## Notes

### Competing Interest Statement

The authors have declared no competing interest.

### Author Declarations

This study only uses openly available human data through the ABCD study.

