## Supplementary material for "Genetic, Hormonal, and Psychosocial Factors Underly Sex Differences in Psychotic-Like Experience Trajectories in Adolescence": Sup. Table 3

**Supplementary Table 3. Two-sample Kolmogorov–Smirnov for PLE symptoms across each year.**

| **Type of PLE** | **Year** | ***D*** | ***p*** | ***p*FDR** |
| --- | --- | --- | --- | --- |
| Hallucination | **Baseline** | **0.0568** | **6.34x10^-6^** | **3.39x10^-5^** |
|  | **Year 1 Follow-up** | **0.0698** | **9.79x10^-9^** | **7.83x10^-8^** |
|  | Year 2 Follow-up | 0.0087 | 0.999 | 0.999 |
|  | Year 3 Follow-up | 0.0294 | 0.067 | -0.107 |
| Delusion | Baseline | 0.0093 | 0.995 | 0.999 |
|  | **Year 1 Follow-up** | **0.0374** | **8.23x10^-3^** | **0.0146** |
|  | Year 2 Follow-up | 0.0087 | 0.998 | 0.999 |
|  | **Year 3 Follow-up** | **0.0487** | **1.82x10^-4^** | **4.86x10^-4^** |
| Disorganized Thoughts and speech | **Baseline** | **0.0478** | **2.6x10^-4^** | **5.90x10^-4^** |
|  | **Year 1 Follow-up** | **0.0517** | **5.62x10^-5^** | **1.80x10^-4^** |
|  | Year 2 Follow-up | 0.0256 | 0.153 | 0.0223 |
|  | **Year 3 Follow-up** | **0.0545** | **1.74x10^-5^** | **6.99x10^-5^** |
| Paranoia | Baseline | 0.0088 | 0.998 | 0.999 |
|  | Year 1 Follow-up | 0.0119 | 0.942 | 0.999 |
|  | Year 2 Follow-up | 0.0471 | 3.34x10^-4^ | 0.223 |
|  | **Year 3 Follow-up** | **0.0922** | **6.39x10^-15^** | **1.02x10^-13^** |

**Significant pFDR values are shown in bold.**
