## Supplementary material for "Genetic, Hormonal, and Psychosocial Factors Underly Sex Differences in Psychotic-Like Experience Trajectories in Adolescence": Sup. Table 5

| Psychosocial Factor | Male Estimate | Female Estimate | Observed Difference | Permutation *p* |
| --- | --- | --- | --- | --- |
| Parent Acceptance (Y1) | -0.09 | -0.44 | 0.35 | 0.0044 |
| Family-related Life events (Y3) | -0.037 | -0.169 | 0.132 | 0.0019 |
| Good life events (Y3) | 0.052 | -0.106 | -0.054 | 0.0156 |
| Overt Aggressor (Y3) | 0.038 | -0.081 | 0.119 | 0.0013 |

**Supplementary Table 5. Permutation test for significant sex difference in psychosocial factors.** Y1 = Year 1 follow-up, Y3 = Year 3 Follow-up.
