## Supplementary material for "Genetic, Hormonal, and Psychosocial Factors Underly Sex Differences in Psychotic-Like Experience Trajectories in Adolescence": Sup. Table 6

| **Report** | **Sex** | **Year** | **β** | **Std. Error** | ***p*** |
| --- | --- | --- | --- | --- | --- |
| Youth | Female | Baseline | 0.52327 | 0.06378 | 3.26x10^-16^ |
|  |  | Follow-up Year 1 | 0.45792 | 0.05072 | <2 x10^-16^ |
|  |  | Follow-up Year 2 | 0.44657 | 0.05026 | <2 x10^-16^ |
|  |  | Follow-up Year 3 | 0.34259 | 0.05705 | 2.07x10^-09^ |
|  | Male | Baseline | 0.93933 | 0.06746 | <2 x10^-16^ |
|  |  | Follow-up Year 1 | 0.18087 | 0.05199 | 5.08 x10^-04^ |
|  |  | Follow-up Year 2 | -0.09558 | 0.07677 | 0.213 |
|  |  | Follow-up Year 3 | 0.32992 | 0.04092 | 9.37 x10^-16^ |
| Parent | Female | Baseline | 0.2258 | 0.05517 | 4.34 x10^-05^ |
|  |  | Follow-up Year 1 | 0.2989 | 0.0492 | 1.35 x10^-09^ |
|  |  | Follow-up Year 2 | 0.44539 | 0.05142 | <2 x10^-16^ |
|  |  | Follow-up Year 3 | 0.40668 | 0.05943 | 8.82 x10^-12^ |
|  | Male | Baseline | 0.41066 | 0.08466 | 1.27 x10^-06^ |
|  |  | Follow-up Year 1 | 0.1256 | 0.0471 | 0.00769 |
|  |  | Follow-up Year 2 | -0.06902 | 0.07062 | 0.328 |
|  |  | Follow-up Year 3 | 0.27043 | 0.04379 | 7.14 x10^-10^ |

**Supplementary Table 6. Linear regression analyses of associations between youth- and parent-reported pubertal status and PLEs.**
