## Supplementary material for "Genetic, Hormonal, and Psychosocial Factors Underly Sex Differences in Psychotic-Like Experience Trajectories in Adolescence": Table 1

| **Variable** | **Pairwise Comparison** | **Mean Difference** | **95% CI** | **Adjusted *p*** |
| --- | --- | --- | --- | --- |
| Age of Pubertal Onset | **Fluctuating vs Decrease** | **-0.164** | **[−0.285, −0.044]** | **0.0026** |
|  | Increase vs Decrease | 0.068 | [-0.153, 0.29] | 0.86 |
|  | Stable vs Decrease | 0.026 | [-0.094, 0.146] | 0.94 |
|  | **Increase vs Fluctuating** | **0.233** | **[0.164, 0.449]** | **0.03** |
|  | **Stable vs Fluctuating** | **0.19** | **[0.081, 0.3]** | **<0.0001** |
|  | Stable vs Increase | -0.042 | [-0.258, 0.174] | 0.96 |
| Age of Menarche | Increase vs Decrease | -0.037 | [-0.26, 0.187] | 0.97 |
|  | Stable vs Decrease | 0.016 | [-0.123, 0.156] | 0.99 |
|  | **Fluctuating vs Decrease** | **-0.26** | **[-0.399, -0.12]** | **<0.0001** |
|  | Stable vs Increase | 0.054 | [-0.158, 0.265] | 0.92 |
|  | **Fluctuating vs Increase** | **-0.223** | **[-0.434, -0.011]** | **0.035** |
|  | **Fluctuating vs Stable** | **-0.276** | **[-0.395, -0.157]** | **<0.0001** |

**Table 1. Tukey HSD post hoc pairwise comparisons of age at pubertal onset and age at menarche across PLE trajectory groups.**
